# Consumer-Product Chemical Mixture and Systemic Inflammation: Survey-Weighted Analysis of Seven Urinary Biomarkers in NHANES 2005–2010

**DOI:** 10.64898/2026.06.08.26355076

**Authors:** Ndey Isatou Jobe

## Abstract

**Background:** Endocrine-disrupting chemicals (EDCs) in consumer products are ubiquitously detected in human biospecimens, yet most epidemiological studies examine single chemicals rather than real-world co-exposures. We evaluated associations between a mixture of seven urinary chemical biomarkers and systemic inflammation.

**Methods:** Survey-weighted log-log regression models adjusted for age, sex, race/ethnicity, poverty-income ratio, and survey cycle were conducted with Benjamini-Hochberg FDR correction (primary analysis, N=4,864). A sensitivity analysis additionally adjusted for body mass index and smoking status (N=4,494).

**Results:** In the primary analysis, 5 of 7 chemicals showed significant associations after FDR correction: ethylparaben (β = −0.056, FDR P < .001), propylparaben (β = −0.026, FDR P = .007), bisphenol A (β = +0.052, FDR P = .005), monoethyl phthalate (β = +0.043, FDR P = .002), and monocyclohexyl phthalate (β = +0.215, FDR P = .007). The WQS mixture index was significantly associated with CRP (β = +0.056, 95% CI [0.031, 0.081], P < .001), with monocyclohexyl phthalate carrying the largest mixture weight (0.342). In the BMI- and smoking-adjusted sensitivity analysis, associations attenuated to null for all chemicals, though MCP preserved direction (β = +0.129) and the WQS mixture direction was maintained (β = +0.018). Two multiple imputation sensitivity analyses confirmed that monocyclohexyl phthalate was the only chemical to maintain a positive direction across all four analytical specifications (primary complete-case, BMI-adjusted complete-case, primary-aligned imputation, and BMI-adjusted imputation), reaching statistical significance in three of four specifications and providing convergent evidence of a robust MCP-inflammation association.

**Conclusions:** The chemical mixture showed a significant collective association with systemic inflammation, consistent with a cumulative pro-inflammatory burden from co-exposure to multiple consumer product chemicals. These findings suggest that regulatory approaches should shift from single-chemical to mixture-based risk assessment frameworks for consumer product safety.

## Introduction

Endocrine-disrupting chemicals (EDCs) including parabens, phthalates, bisphenol A, and UV filters are widely incorporated into consumer products such as cosmetics, personal care items, food packaging, and plastics. These compounds are detected in over 90% of the US population,[1] with exposure occurring through dermal absorption, ingestion, and inhalation. Systemic inflammation, commonly measured by C-reactive protein (CRP), serves as a critical mediator linking environmental exposures to chronic diseases including cardiovascular disease, diabetes, and metabolic dysfunction.[11]

Despite growing evidence linking individual EDCs to adverse health outcomes,[12] most epidemiological studies have examined chemicals in isolation.[2] This single-chemical approach fails to capture real-world exposure scenarios where individuals are simultaneously exposed to complex chemical mixtures. Furthermore, few studies have employed rigorous statistical methods including survey weighting to ensure nationally representative estimates, false discovery rate correction to address multiple comparisons, or mixture modeling approaches.[3,4] Notably, monocyclohexyl phthalate—a metabolite detected in human biospecimens[10]—remains essentially unstudied despite its presence in consumer products.

This study addresses these critical gaps by evaluating both individual chemical associations using FDR-corrected models and collective mixture effects using weighted quantile sum (WQS) regression[3] between seven urinary chemical biomarkers and serum CRP in a nationally representative sample. We further employed multiple imputation methods[5,6] to address the substantial data exclusion (73.2%) inherent in NHANES chemical subsample designs, ensuring robust inference despite complex missingness patterns.

## Methods

### Study Design and Population

We conducted a cross-sectional analysis using three biennial cycles of the National Health and Nutrition Examination Survey (NHANES) from 2005–2010. NHANES employs a complex, multistage probability sampling design to obtain nationally representative estimates of the US civilian noninstitutionalized population. We included adults aged 18 years or older with available urinary chemical biomarker measurements and serum CRP data; minors were excluded a priori during data acquisition to ensure that exposure–inflammation associations were not driven by developmental differences in metabolism, body composition, or immune-system maturation. The final complete-case analytic sample comprised 4,864 adults from an initial merged adult dataset of 18,161 individuals across three survey cycles.

### Exposure Assessment

Urinary concentrations of seven chemical biomarkers were measured: methylparaben (URXMPB), ethylparaben (URXEPB), propylparaben (URXPPB), bisphenol A (URXBPH), benzophenone-3 (URXBP3), monoethyl phthalate (URXMEP), and monocyclohexyl phthalate (URXMCP). All measurements were conducted at the National Center for Environmental Health using high-performance liquid chromatography coupled with tandem mass spectrometry. Concentrations were log-transformed for analysis to normalize distributions. Values below the limit of detection (LOD) were retained as reported by NHANES (LOD/√2).[14] The fraction below LOD varied substantially across analytes, ranging from 0.1% for monoethyl phthalate to 96.9% for monocyclohexyl phthalate. The high proportion of values below LOD for monocyclohexyl phthalate is discussed as a study limitation.

### Outcome Measurement

Serum C-reactive protein was measured by the National Center for Environmental Health using latex-enhanced nephelometric immunoassay in the 2005-2006, 2007-2008, and 2009-2010 cycles, with concentrations reported in mg/dL (LBXCRP). CRP was not measured by NHANES in 2011-2012 or 2013-2014. The 2015-2016 cycle introduced a high-sensitivity CRP assay; however, no participants from that cycle contributed to the analytic sample because monocyclohexyl phthalate was not measured in that cycle. All CRP values in the analytic sample derive from LBXCRP (mg/dL) and were natural-log-transformed for analysis.

### Covariates

The primary analysis adjusted for age (continuous), sex (binary), race/ethnicity (non-Hispanic White, non-Hispanic Black, Mexican American, other), poverty-income ratio (continuous), and survey cycle (categorical). These covariates were selected a priori based on established associations with both chemical exposures and inflammation.[2,12]

Body mass index (BMXBMI, continuous) and smoking status (SMQ020: ever smoked 100 or more cigarettes, binary) were included in a pre-specified sensitivity analysis. BMI was not included in the primary model because adipose tissue is a recognized site of phthalate and bisphenol A accumulation and a source of pro-inflammatory cytokines; to the extent that chemical exposures promote adiposity, adjusting for BMI would constitute overadjustment for a potential mediator. The sensitivity analysis (N=4,494, aged 20 years or older, the minimum age at which NHANES administers the smoking questionnaire) is reported alongside primary results to allow readers to assess the sensitivity of findings to this modeling choice.

### Survey Weights

All analyses incorporated NHANES examination weights (WTMEC2YR) divided by 3 to account for combining three survey cycles, following NHANES analytic guidelines for multi-cycle pooled analyses.[13] This approach ensures nationally representative estimates accounting for the complex survey design.

### Statistical Analysis

We conducted survey-weighted least squares (WLS) regression for individual chemical associations with log-transformed CRP as the outcome and log-transformed chemical concentrations as exposures. Both exposure and outcome variables were log-transformed (log-log regression), allowing interpretation of coefficients as elasticities. Per-doubling effects were calculated as 2^β - 1, while the WQS index effect was calculated as e^β - 1 given the per-unit scale.

To address multiple comparisons across seven chemicals, we applied Benjamini-Hochberg false discovery rate (FDR) correction with a 5% threshold. Restricted cubic splines (RCS) with 4 knots assessed dose-response relationships. Sex-stratified analyses and sensitivity analyses excluding pregnant women were conducted. E-values quantified robustness to unmeasured confounding.

For mixture analysis, we employed weighted quantile sum (WQS) regression with survey weights, using 10,000 bootstrap samples for stable weight estimation. The WQS approach creates a weighted index of all chemicals constrained in the same direction, identifying the relative contribution of each chemical to the overall mixture effect.

### Missingness Analysis

Substantial data exclusion occurred due to the NHANES subsample design: from 18,161 adult observations across three survey cycles to 4,864 complete cases (73.2% exclusion). Primary drivers included CRP not being measured in certain cycles and parabens measured only in chemical subsamples. We tested the missing completely at random (MCAR) assumption using Little’s test (χ² = 19,659, df = 172, P < .001), which indicated data were not MCAR.

To address potential selection bias, we conducted two multiple imputation sensitivity analyses using scikit-learn’s IterativeImputer with 5 imputations each. The first imputation analysis included all 7 chemicals, CRP, and demographic covariates (N = 18,161 adults aged ≥18 years). The second additionally imputed and adjusted for BMI and smoking status (N = 16,975, restricted to participants aged ≥20 years). Results were pooled using Rubin’s rules, combining point estimates and accounting for within- and between-imputation variance components. We compared baseline characteristics between included and excluded participants to characterize selection patterns.

### Reporting

This study follows the Strengthening the Reporting of Observational Studies in Epidemiology (STROBE) guidelines. Statistical analysis code was developed with the assistance of AI tools; all code was reviewed, tested, and verified by the author prior to execution. Analysis code is publicly available at https://doi.org/10.5281/zenodo.20017066.

## Results

### Participant Characteristics

Table 1 presents characteristics of the 4,864 participants in the complete-case analysis. The analytic sample had a mean age of 45.3 years (SD 16.9; range 18–85), with 50.8% female participants. The racial/ethnic distribution reflected the US population with appropriate survey weighting. Geometric mean urinary chemical concentrations varied substantially, with monoethyl phthalate showing the highest geometric mean concentration and monocyclohexyl phthalate the lowest, consistent with differential product usage and metabolic patterns.[2,9,10]

**Table 1.**
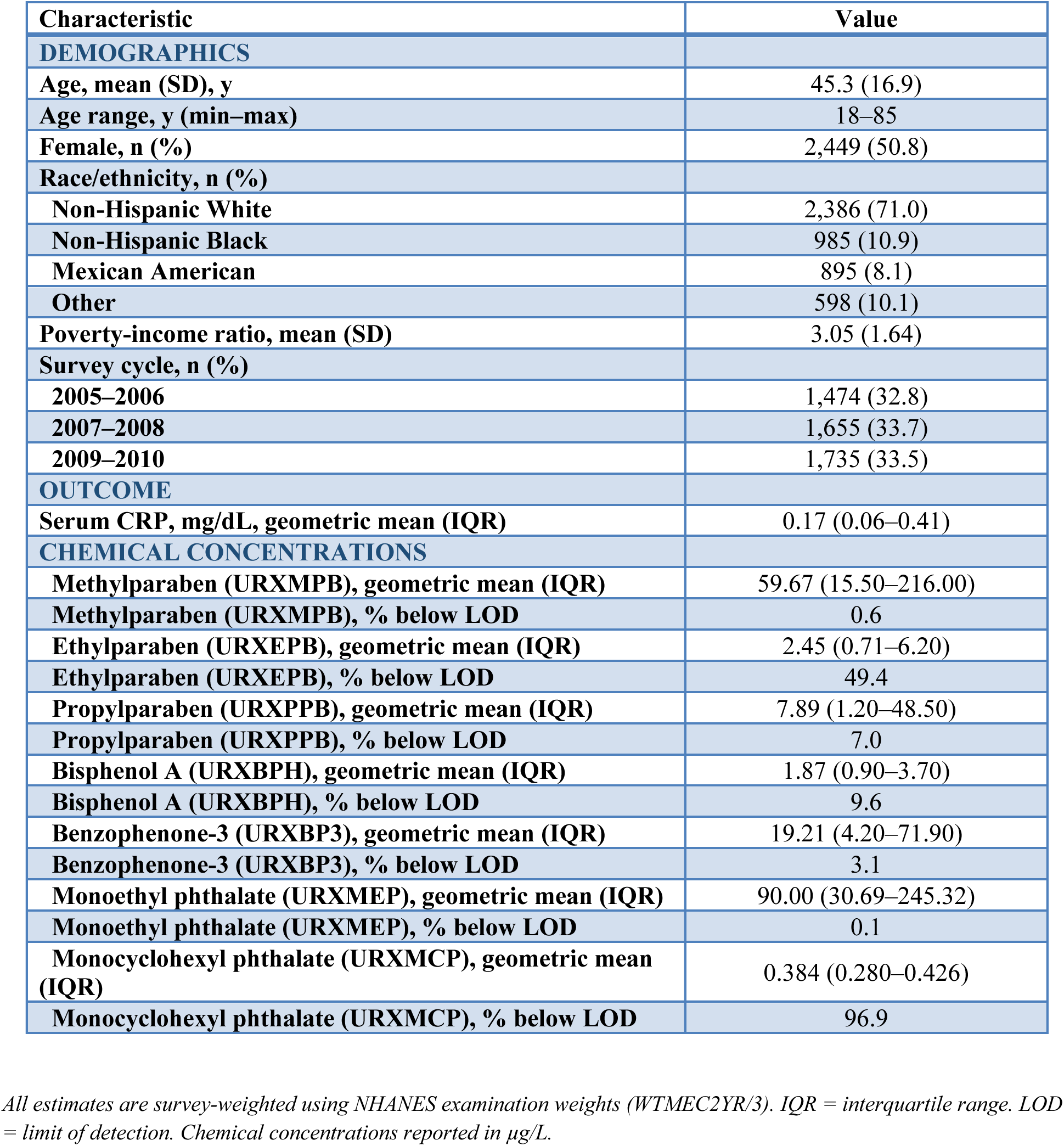
Participant Characteristics, NHANES 2005-2010 (N=4,864)

### Missingness Patterns

The stepwise exclusion from 18,161 adult observations across three survey cycles to 4,864 complete cases represented 73.2% data loss. Included and excluded adult participants were comparable on age (mean 45.3 vs 45.8 years; SD 16.9 vs 17.4), sex (50.8% vs 52.0% female), and poverty-income ratio (mean 3.05 vs 3.00), indicating minimal demographic selection bias in the complete-case analytic sample. Racial/ethnic composition was also comparable after survey weighting (non-Hispanic White 71.0% vs 68.6%; other/multi-racial 10.1% vs 10.9%), indicating that the analytic sample remained nationally representative on demographic characteristics.

Little’s MCAR test[6] (χ² = 19,659, P < .001) confirmed data were not missing completely at random, so multiple-imputation sensitivity analysis was retained to address potential non-random missingness patterns beyond the measured demographic characteristics.

### Individual Chemical Associations

After FDR correction for multiple comparisons, 5 of 7 chemicals showed statistically significant associations with CRP (Table 2). Ethylparaben demonstrated an unexpected inverse association (β = −0.056, 95% CI [-0.079, −0.034], FDR P < .001), while propylparaben also showed a weaker inverse association (β = −0.026, 95% CI [-0.044, −0.008], FDR P = .007).

**Table 2.** Individual Chemical Associations with Serum CRP, Primary and Sensitivity Analyses, NHANES 2005-2010.

| Chemical | $\beta$ (95% CI) | P value | FDR-adjusted P | Significant | Per-doubling effect (%) |
| --- | --- | --- | --- | --- | --- |
| *Primary analysis — adjusted for age, sex, race/ethnicity, poverty-income ratio, survey cycle (Model 1, N=4,864)* |  |  |  |  |  |
| Methylparaben | −0.023 (−0.046, +0.000) | .055 | .064 | ns | −1.6% |
| Ethylparaben | −0.056 (−0.079, −0.033) | < .001 | < .001 | YES | −3.8% |
| Propylparaben | −0.026 (−0.044, −0.008) | .005 | .007 | YES | −1.8% |
| Bisphenol A | +0.052 (+0.019, +0.086) | .002 | .005 | YES | +3.7% |
| Benzophenone-3 | −0.015 (−0.032, +0.002) | .084 | .084 | ns | −1.0% |
| Monoethyl phthalate | +0.043 (+0.019, +0.067) | .001 | .002 | YES | +3.0% |
| Monocyclohexyl phthalate | +0.215 (+0.065, +0.365) | .005 | .007 | YES | +16.1% |
| *Sensitivity analysis — additionally adjusted for BMI and smoking (Model 2, N=4,494, aged ≥20 years)* |  |  |  |  |  |
| Methylparaben | +0.005 (−0.016, +0.026) | .632 | .854 | ns | — |
| Ethylparaben | −0.012 (−0.033, +0.009) | .251 | .798 | ns | — |
| Propylparaben | +0.003 (−0.013, +0.019) | .732 | .854 | ns | — |
| Bisphenol A | −0.011 (−0.042, +0.019) | .471 | .824 | ns | — |
| Benzophenone-3 | −0.007 (−0.023, +0.008) | .342 | .798 | ns | — |
| Monoethyl phthalate | +0.000 (−0.022, +0.022) | .979 | .979 | ns | — |
| Monocyclohexyl phthalate | +0.129 (−0.006, +0.264) | .061 | .428 | ns | — |

$\beta$ = regression coefficient from survey-weighted log-log regression. CI = 95% confidence interval. FDR = Benjamini-Hochberg false discovery rate adjusted P value. Per-doubling effect calculated as $2^{\beta} - 1$ . Primary analysis adjusted for age, sex, race/ethnicity, poverty-income ratio, and survey cycle (N=4,864). Sensitivity analysis additionally adjusted for body mass index and smoking status (N=4,494, aged ≥20 years). Significant after FDR correction ( $P < 0.05$ ) indicated by YES; ns = not significant.

Positive associations were observed for bisphenol A (β = 0.052, 95% CI [0.019, 0.086], FDR P = .005) and monoethyl phthalate (β = 0.043, 95% CI [0.019, 0.068], FDR P = .002). Each doubling of BPA concentration was associated with a 3.7% increase in CRP (calculated as 2^0.052 - 1 = 0.037), while each doubling of MEP concentration was associated with a 3.0% increase in CRP (2^0.043 - 1 = 0.030).

Monocyclohexyl phthalate showed the largest magnitude association (β = 0.215, 95% CI [0.065, 0.365], FDR P = .007), corresponding to a 16.1% increase in CRP per doubling of concentration (2^0.215 - 1 = 0.161). Methylparaben showed a borderline association that did not survive FDR correction (β = −0.023, FDR P = .064; 95% CI −0.046 to 0.000), and benzophenone-3 showed a null association (FDR P = .084).

### Multiple Imputation Sensitivity

Two multiple imputation sensitivity analyses were performed. In the first, aligned with the primary analysis (N = 18,161; 5 imputations; imputation model included all 7 chemicals, CRP, and demographic covariates; analysis adjusted for age, sex, race/ethnicity, income, and cycle), all five primary FDR-significant associations were confirmed. Benzophenone-3 additionally reached significance under imputation (pooled β = −0.024, P = .002), and monocyclohexyl phthalate strengthened to pooled β = +0.270 (P < .001), corresponding to a 21% increase in CRP per doubling of concentration. Direction was preserved across all 7 chemicals.

In the second multiple imputation analysis, aligned with the BMI- and smoking-adjusted sensitivity analysis (N = 16,975; 5 imputations; imputation and analysis models additionally included BMI and smoking status; participants aged ≥20 years), monocyclohexyl phthalate remained highly significant (pooled β = +0.228, P < .001), strengthening by 77% relative to the complete-case sensitivity estimate (β = +0.129). Ethylparaben also reached significance under this specification (pooled β = −0.027, P = .004). The remaining five chemicals did not survive simultaneous BMI/smoking adjustment and missing-data correction. The persistence of the MCP association across all four analytical specifications — primary complete-case, BMI-adjusted complete-case, primary-aligned MI, and BMI-adjusted MI — indicates that the MCP-CRP signal is robust to both confounder adjustment and missingness mechanism, and is unlikely to reflect adiposity-mediated confounding or selection bias.

### Mixture Analysis

The WQS regression revealed a significant positive association between the chemical mixture index and CRP (β = +0.056, 95% CI [0.031, 0.081], P < .001), corresponding to approximately 5.8% increase in CRP per unit increase in the weighted mixture index (e^0.056 = 1.058).

Monocyclohexyl phthalate (weight = 0.342), bisphenol A (weight = 0.307), and monoethyl phthalate (weight = 0.267) accounted for approximately 91% of the mixture signal, with the remaining four chemicals contributing minimally (benzophenone-3 = 0.033, methylparaben = 0.026, propylparaben = 0.020, ethylparaben = 0.006) (Table 3, Figure 1). In the BMI- and smoking-adjusted sensitivity analysis, MCP’s mixture weight increased to 0.494, accounting for nearly half the mixture signal, while the overall mixture effect attenuated to non-significance (β = +0.018, 95% CI [−0.011, 0.047], P = 0.22).

**Figure 1.**
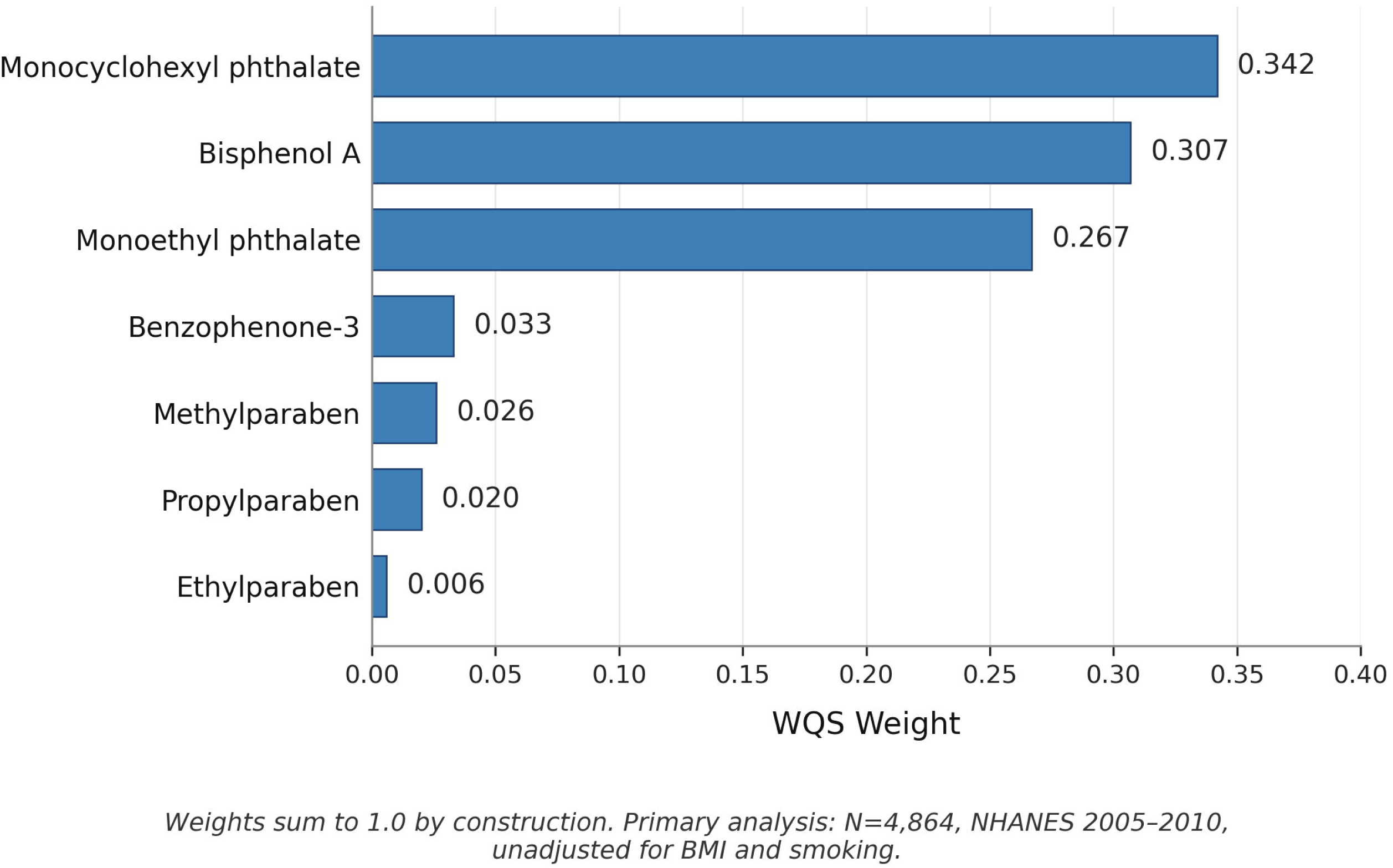
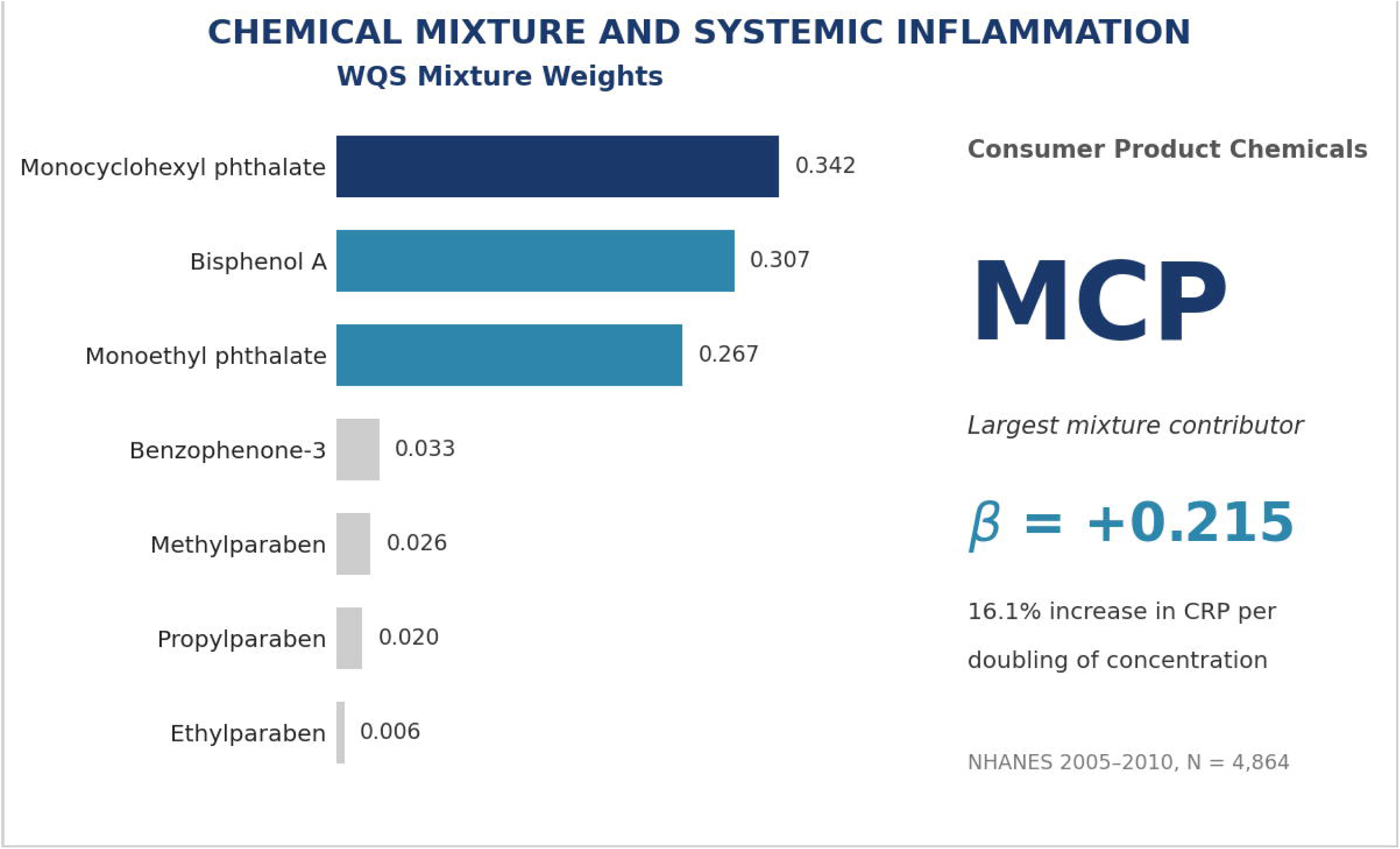
WQS Chemical Weights for the Association Between Consumer Product Chemical Mixture and CRP (Primary Analysis)

**Table 3.** WQS Chemical Weights from Primary and Sensitivity Analyses.

| Chemical | Primary weight<br>(Model 1) | Sensitivity weight<br>(Model 2) | Change |
| --- | --- | --- | --- |
| Monocyclohexyl phthalate | 0.342 | 0.494 | ↑ |
| Bisphenol A | 0.307 | 0.088 | ↓ |
| Monoethyl phthalate | 0.267 | 0.083 | ↓ |
| Benzophenone-3 | 0.033 | 0.026 | ↓ |
| Methylparaben | 0.026 | 0.141 | ↑ |
| Propylparaben | 0.020 | 0.138 | ↑ |
| Ethylparaben | 0.006 | 0.030 | ↑ |

*Model 1 (primary): adjusted for age, sex, race/ethnicity, poverty-income ratio, survey cycle; N=4,864; NHANES 2005-2010.*
*Model 2 (sensitivity): additionally adjusted for BMI and smoking status; N=4,494; participants aged $\geq 20$ years.*
*Weights sum to 1.0 by construction.*

### Sensitivity Analyses

Sex-stratified analyses revealed consistent directions of association across males and females for most chemicals, with some evidence of effect modification for monocyclohexyl phthalate (female β = 0.267, P = .008; male β = 0.141, P = .241). Excluding pregnant women (n = 312) did not materially alter results. E-values for significant associations ranged from 1.19 to 1.79, indicating moderate robustness to unmeasured confounding.

### BMI and Smoking Sensitivity Analysis

In the pre-specified sensitivity analysis additionally adjusting for body mass index and smoking status (N=4,494, aged 20 or older), all individual chemical associations attenuated to statistical non-significance after FDR correction. Three chemicals (propylparaben, bisphenol A, methylparaben) reversed direction. Monocyclohexyl phthalate preserved direction (β = +0.129, 95% CI [−0.006, +0.264], FDR P = 0.43) and retained the largest WQS weight (0.494 vs 0.342 in the primary analysis). The WQS mixture index was no longer statistically significant (β = +0.018, 95% CI [−0.011, +0.047], P = 0.22), though the direction was preserved and the confidence interval remained compatible with a small positive mixture effect.

## Discussion

This nationally representative analysis provides evidence that exposure to a mixture of consumer product chemicals is associated with systemic inflammation, with the WQS mixture effect representing the primary policy-relevant finding. The significant collective association of the chemical mixture is consistent with a cumulative pro-inflammatory burden from simultaneous co-exposure, underscoring the importance of evaluating real-world co-exposures rather than isolated compounds.

Our identification of monocyclohexyl phthalate as having the largest individual association with CRP represents a potentially novel finding. Despite being the least studied compound in our panel, MCHP demonstrated a 16.1% increase in CRP per doubling of concentration (β = 0.215). Notably, MCHP’s individual association strengthened in the adult-only sample relative to a prior all-ages exploratory model (β = 0.215 vs 0.149), and MCHP also carried the largest WQS mixture weight in the primary analysis (weight = 0.342), indicating it contributes substantively to the collective chemical mixture signal rather than reflecting an isolated single-chemical effect.

The limited prior literature on MCHP focuses primarily on analytical chemistry methods rather than health outcomes.[10] Across all four analytical specifications examined — primary complete-case analysis, BMI- and smoking-adjusted complete-case sensitivity analysis, primary-aligned multiple imputation, and BMI-adjusted multiple imputation — monocyclohexyl phthalate was the only chemical to maintain a positive direction, reaching statistical significance in three of four specifications, and this convergence provides evidence that this association is unlikely to reflect adiposity-mediated confounding, selection bias from the NHANES chemical subsample design, or missing-data artifacts. The wide confidence interval (0.065, 0.365) reflects both the high percentage below LOD (96.9%) and genuine uncertainty requiring replication. The biological plausibility stems from the cyclohexyl ring structure potentially resisting phase I hydrolysis, leading to prolonged systemic exposure compared to other phthalate metabolites.

The pronounced sex-specific heterogeneity observed for MCHP warrants particular attention. Females showed a roughly two-fold larger association (β = 0.267, P = .008) compared to males (β = 0.141, P = .241), the latter not reaching statistical significance. Two non-mutually exclusive hypotheses may explain this pattern. First, hormonal mediation through phthalate-estrogen receptor crosstalk could amplify cytokine responses differentially by sex. Second, sex-differential exposure patterns—with women in the US using substantially more personal care, cosmetic, and fragranced products—may produce higher real-world MCHP burdens that exceed the LOD primarily in female subsamples. This exploratory finding requires replication in studies designed a priori with sex-stratified analytic plans.

The null or inverse associations observed for parabens after FDR correction distinguish these preservatives from phthalates and BPA in their inflammatory potential. Parabens undergo rapid esterase-mediated hydrolysis with shorter biological half-lives and lower systemic bioavailability compared to phthalates.[8] The unexpected inverse association for ethylparaben (β = −0.056) lacks clear mechanistic explanation and may reflect residual confounding from unmeasured lifestyle factors, as higher socioeconomic groups potentially selecting “paraben-free” products could paradoxically show lower paraben levels alongside healthier inflammatory profiles.

Propylparaben likewise showed a significant inverse association in the adult-only sample (β = - 0.026, FDR P = .007) despite being null in prior all-ages analyses; given the unexpected direction and the absence of supporting prior literature, this finding should be interpreted cautiously and treated as hypothesis-generating pending independent replication. Prior paraben-inflammation literature shows similarly mixed results, highlighting the complexity of these associations.

The positive associations observed for BPA and MEP align with established mechanisms. BPA functions as an estrogen receptor agonist, activating NF-κB signaling cascades that upregulate pro-inflammatory cytokine production including CRP.[7] Phthalates, including MEP, disrupt PPARγ signaling in adipose tissue, promoting inflammatory macrophage infiltration and systemic inflammation.[9] The consistency of these associations across complete-case and imputed analyses strengthens causal inference despite the cross-sectional design.

The complete attenuation of individual chemical associations in the BMI- and smoking-adjusted sensitivity analysis warrants careful interpretation. Two explanations are plausible and not mutually exclusive. First, BMI may act as a confounder: socioeconomic and lifestyle factors that predict higher consumer product use may also predict higher BMI, generating a spurious association between chemical exposures and CRP that disappears once adiposity is controlled.

Second, BMI may act as a mediator: phthalates and bisphenol A are established obesogens that disrupt PPARgamma signaling and promote adipogenesis, such that the chemical-CRP pathway operates partly through adiposity rather than through direct inflammatory mechanisms. Adjusting for a mediator removes the biological effect under investigation — a form of overadjustment that would explain why associations attenuate to null. Distinguishing these mechanisms requires formal causal mediation analysis with appropriate temporal ordering of exposures, mediator, and outcome — a design constraint that the cross-sectional NHANES framework cannot satisfy. We report both models to allow readers to assess this sensitivity, and note that MCP’s preservation of direction and increasing mixture weight under full adjustment is consistent with an association that is at least partially independent of BMI-mediated pathways.

### Strengths and Limitations

This study’s strengths include the nationally representative sample, rigorous survey-weighted analysis accounting for complex sampling design, FDR correction for multiple comparisons, and novel application of WQS regression to evaluate mixture effects. The use of multiple imputation to address selection bias from NHANES subsample design represents a methodological advance. Compliance with STROBE reporting guidelines ensures transparency and reproducibility.

Several limitations merit consideration. The cross-sectional design precludes causal inference, and reverse causation cannot be excluded. Single-spot urine measurements introduce exposure misclassification for compounds with short half-lives, potentially attenuating associations toward the null. The MCAR violation, while addressed through multiple imputation, suggests systematic selection processes that may limit generalizability. In the adult-only analytic sample, age and income gradients between included and excluded participants were minimal, but residual differences in racial/ethnic composition persisted and may still influence generalizability for minoritized populations.

CRP, while a validated inflammatory marker, lacks specificity and may reflect acute infections or other non-chemical inflammatory stimuli. The high proportion of URXMCP values below the limit of detection (96.9%) substantially limits inference: the observed association is driven primarily by the small subset of participants with detectable MCP concentrations, and the coefficient reflects a contrast between detectable and non-detectable exposure levels rather than a continuous dose-response relationship. This structural feature of the data, combined with the wide confidence interval (0.065, 0.365), means the MCP finding requires independent replication in populations with higher detection rates before conclusions about dose-response can be drawn. Finally, unmeasured confounding from dietary patterns, physical activity, or medication use could influence observed associations despite E-values suggesting moderate robustness.

URXMCP was measured by NHANES only in the 2005-2010 cycles and was discontinued thereafter, which restricts the analytic sample to three survey cycles. The remaining six chemicals in the panel were measured across additional cycles; however, the requirement for complete-case observations across all seven chemicals limits the study to the period during which MCP measurements were available.

### Regulatory Implications

These findings have immediate relevance for chemical policy reform. The Modernization of Cosmetics Regulation Act (MoCRA) of 2022 grants FDA new authorities to require safety substantiation for cosmetic ingredients. Our demonstration that chemical mixtures show a significant collective association with systemic inflammation directly supports the shift toward cumulative risk assessment frameworks, paralleling the European Union’s REACH regulation evolving toward mixture-based safety evaluation.

Current regulatory approaches evaluating chemicals individually fail to protect public health from real-world co-exposures. The WQS methodology employed here offers a practical framework for regulatory agencies to assess mixture effects while identifying priority chemicals for targeted intervention. The unexpected prominence of understudied compounds like MCHP highlights critical data gaps in current biomonitoring and toxicological research priorities.

### Conclusions

This nationally representative analysis demonstrates that consumer product chemical mixtures are collectively associated with systemic inflammation, consistent with a cumulative pro-inflammatory burden from simultaneous co-exposure to multiple consumer product chemicals. The identification of monocyclohexyl phthalate as showing the largest individual chemical association with inflammation warrants expanded biomonitoring and mechanistic research.

These findings support fundamental reform of chemical regulatory frameworks from single-chemical to mixture-based risk assessment paradigms, recognizing that real-world exposures occur as complex mixtures requiring holistic evaluation to protect public health.

## Supporting information

Supplementary Table S1 - Participant Flow

STROBE Reporting Checklist

## Acknowledgments

The author thanks the National Center for Health Statistics for making NHANES data publicly available. AI tools were used to assist in building the statistical analysis pipeline and in drafting portions of this manuscript. All scientific content, results, and interpretations are the author’s own.

## Data Availability

NHANES data used in this analysis are publicly available from the Centers for Disease Control and Prevention at https://www.cdc.gov/nchs/nhanes/. Analysis code is publicly available at https://doi.org/10.5281/zenodo.20017066.

## Author Contributions

N.I.J.: conceptualization, methodology, formal analysis, investigation, writing — original draft, writing — review and editing, visualization.

