## Supplementary Table S1 - Participant Flow for "Consumer-Product Chemical Mixture and Systemic Inflammation: Survey-Weighted Analysis of Seven Urinary Biomarkers in NHANES 2005–2010"

**Supplementary Table S1. Participant Inclusion and Exclusion Flow, NHANES 2005-2010**

| **Step** | **Criteria** | **N remaining** | **N excluded this step** | **% excluded this step** | **Cumulative % excluded** |
| --- | --- | --- | --- | --- | --- |
| 0 | Full NHANES parquet downloaded (cycles 2005-2016); analysis restricted to cycles 2005-2010 | 36,287 | — | — | 0.00% |
| 1 | Restrict to NHANES cycles 2005-2010 (SDDSRVYR ∈ {4, 5, 6}) | 18,318 | 17,969 | 49.52% | 49.52% |
| 2 | Restrict to adults aged ≥ 18 years (RIDAGEYR ≥ 18) | 18,161 | 157 | 0.86% | 49.95% |
| 3 | Exclude missing serum CRP (LBXCRP not null) | 16,467 | 1,694 | 9.33% | 54.62% |
| 4 | Exclude missing any of 7 urinary chemical biomarkers (URXMPB, URXEPB, URXPPB, URXBPH, URXBP3, URXMEP, URXMCP) | 5,262 | 11,205 | 68.05% | 85.50% |
| 5 | Exclude missing any covariate (age, sex, race/ethnicity, poverty-income ratio, survey cycle) | 4,864 | 398 | 7.56% | 86.60% |
| **6** | **Exclude participants with zero or missing examination survey weight (WTMEC2YR > 0)** | **4,864** | **0** | **0.00%** | **86.60%** |

*NHANES = National Health and Nutrition Examination Survey. CRP = C-reactive protein. The analytic sample of 4,864 adults includes all participants with complete data on serum CRP, all seven urinary chemical biomarkers, and demographic covariates in NHANES cycles 2005-2006, 2007-2008, and 2009-2010.*
