## Supplementary material for "Consumer-Product Chemical Mixture and Systemic Inflammation: Survey-Weighted Analysis of Seven Urinary Biomarkers in NHANES 2005–2010": STROBE Reporting Checklist

### STROBE Statement — Cross-Sectional Studies Checklist

**Companion to:** JOBE_ChemicalMixtures_NHANES_FINAL.docx

| **Item #** | **STROBE requirement** | **Where addressed in manuscript** |
| --- | --- | --- |
| 1(a) | Indicate the study design with a commonly used term in the title or the abstract | Title: "Survey-Weighted Analysis... NHANES 2005–2010"; Abstract Methods sentence 1 |
| 1(b) | Provide in the abstract an informative and balanced summary of what was done and what was found | Abstract (Background, Methods, Results, Conclusions) |
| 2 | Background/rationale: explain the scientific background and rationale for the investigation | Introduction, paragraphs 1–2 |
| 3 | Objectives: state specific objectives, including any prespecified hypotheses | Introduction, paragraph 3 |
| 4 | Study design: present key elements of study design early in the paper | Methods §Study Design and Population, sentence 1 ("cross-sectional analysis using three biennial cycles…") |
| 5 | Setting: describe the setting, locations, and relevant dates, including periods of recruitment, exposure, follow-up, and data collection | Methods §Study Design and Population (NHANES 2005–2010, US civilian noninstitutionalized population) |
| 6(a) | Participants: give the eligibility criteria, and the sources and methods of selection of participants | Methods §Study Design and Population (adults ≥18 with urinary biomarkers and CRP); Results §Participant Characteristics |
| 7 | Variables: clearly define all outcomes, exposures, predictors, potential confounders, and effect modifiers; give diagnostic criteria, if applicable | Methods §Exposure Assessment, §Outcome Measurement, §Covariates |
| 8 | Data sources/measurement: for each variable of interest, give sources of data and details of methods of assessment (measurement); describe comparability of assessment methods if there is more than one group | Methods §Exposure Assessment (HPLC-MS/MS at NCEH); §Outcome Measurement (high-sensitivity nephelometry, HSCRP) |
| 9 | Bias: describe any efforts to address potential sources of bias | Methods §Survey Weights (WTMEC2YR/3); §Statistical Analysis (FDR correction, E-values); §Missingness Analysis (Little's MCAR test, multiple imputation); Results §Missingness Patterns |
| 10 | Study size: explain how the study size was arrived at | Methods §Study Design and Population (final N = 4,864 from merged adult dataset of 18,161 after complete-case filtering); Results §Missingness Patterns (stepwise exclusion) |
| 11 | Quantitative variables: explain how quantitative variables were handled in the analyses; if applicable, describe which groupings were chosen and why | Methods §Statistical Analysis (log-log regression; restricted cubic splines with 4 knots; WQS quantile-based index) |
| 12(a) | Statistical methods: describe all statistical methods, including those used to control for confounding | Methods §Statistical Analysis (WLS regression, log-log specification, covariate set) |
| 12(b) | Describe any methods used to examine subgroups and interactions | Methods §Statistical Analysis (sex-stratified analyses; sensitivity analysis excluding pregnant women); Results §Sensitivity Analyses |
| 12(c) | Explain how missing data were addressed | Methods §Missingness Analysis (Little's MCAR; multiple imputation N=18,161 and N=16,975, Rubin's rules); Results §Multiple Imputation Sensitivity |
| 12(d) | If applicable, describe analytical methods taking account of sampling strategy | Methods §Survey Weights (WTMEC2YR/3, multi-cycle pooling per NHANES analytic guidelines) |
| 12(e) | Describe any sensitivity analyses | Methods §Statistical Analysis & §Missingness Analysis; Results §Multiple Imputation Sensitivity, §Sensitivity Analyses (sex-stratified, no-pregnant-women, E-values) |
| 13(a) | Participants: report numbers of individuals at each stage of study — eg numbers potentially eligible, examined for eligibility, confirmed eligible, included in the study, completing follow-up, and analysed | Results §Missingness Patterns (18,161 → 4,864, with stepwise exclusion); Methods §Study Design and Population |
| 13(b) | Give reasons for non-participation at each stage | Results §Missingness Patterns (subsample design; CRP not in all cycles; parabens in chemical subsamples) |
| 13(c) | Consider use of a flow diagram | Supplementary Table S1 provides a stepwise participant inclusion and exclusion flow table (18,161 → 4,864 with N excluded and % excluded at each step) |
| 14(a) | Descriptive data: give characteristics of study participants (eg demographic, clinical, social) and information on exposures and potential confounders | Results §Participant Characteristics (Table 1); Methods §Covariates |
| 14(b) | Indicate number of participants with missing data for each variable of interest | Results §Missingness Patterns (stepwise table-style narrative: ln_LBXCRP, ln_URXMPB, ln_URXBP3, age, income exclusions) |
| 15 | Outcome data: report numbers of outcome events or summary measures | Results §Individual Chemical Associations (Table 2 with β, 95% CI, FDR P for each chemical); §Mixture Analysis (Table 3, Figure 1) |
| 16(a) | Main results: give unadjusted estimates and, if applicable, confounder-adjusted estimates and their precision (eg, 95% confidence interval); make clear which confounders were adjusted for and why they were included | Results §Individual Chemical Associations (adjusted β with 95% CI for all 7 chemicals); Methods §Covariates lists adjusted variables |
| 16(b) | Report category boundaries when continuous variables were categorized | Not applicable — exposures and outcome modeled as continuous (log-transformed); WQS uses internal quantile binning per Carrico et al. [3] |
| 16(c) | If relevant, translate estimates of relative risk into absolute risk for a meaningful time period | Not applicable — cross-sectional design; per-doubling and per-unit-WQS percentage changes in CRP reported instead (Results §Individual Chemical Associations) |
| 17 | Other analyses: report other analyses done — eg analyses of subgroups and interactions, and sensitivity analyses | Results §Multiple Imputation Sensitivity; §Sensitivity Analyses (sex-stratified; no-pregnant-women; E-values) |
| 18 | Key results: summarise key results with reference to study objectives | Discussion, paragraph 1 |
| 19 | Limitations: discuss limitations of the study, taking into account sources of potential bias or imprecision; discuss both direction and magnitude of any potential bias | Discussion §Strengths and Limitations |
| 20 | Interpretation: give a cautious overall interpretation of results considering objectives, limitations, multiplicity of analyses, results from similar studies, and other relevant evidence | Discussion (paragraphs on MCHP, age-dependent mixture, sex heterogeneity, parabens, BPA/MEP mechanisms) |
| 21 | Generalisability: discuss the generalisability (external validity) of the study results | Discussion §Strengths and Limitations (selection bias on race/ethnicity); §Regulatory Implications |
| 22 | Funding: give the source of funding and the role of the funders for the present study and, if applicable, for the original study on which the present article is based | Funding section ("This research received no external funding") |

**Source:** von Elm E, et al. The Strengthening the Reporting of Observational Studies in Epidemiology (STROBE) statement: guidelines for reporting observational studies. Bull World Health Organ. 2007;85(11):867–872. (Cross-sectional studies version, items 1–22.)
